# A multi-dimensional framework for establishing and managing a genomic newborn screening program

**DOI:** 10.1101/2025.06.17.25329471

**Authors:** Elena Schnabel-Besson, Nicola Dikow, Karla Alex, Ulrike Mütze, Hannah Straub, Elena Sophia Doll, Julia Mahal, Heiko Brennenstuhl, Carlotta Julia Mayer, Lars Neth, Tobias Hagedorn, Henriette Högl, Sascha Settegast, Beate Ditzen, Ralf Müller-Terpitz, Stefan Kölker, Christian P. Schaaf, Eva Winkler

## Abstract

**Purpose:** Newborn screening (NBS) is an effective measure of secondary prevention. The application of genomic sequencing in population-based screening would enable further expansions of the NBS disease panel and a genomic NBS (gNBS). The selection of NBS target diseases is still based on the Wilson and Jungner screening criteria from 1968, which are considered incomplete, rendering the necessity of developing new criteria.

**Methods:** The present work aims to establish a multi-dimensional framework for future gNBS programs. An interdisciplinary expert panel comprising researchers from pediatric and adolescent medicine, human genetics, ethics, medical psychology, law, and patient representatives used a nominal group technique-like multi-stage consensus process to define criteria for gNBS, considering ethical, legal, and social implications, medical aspects, and patient perspectives.

**Results:** Overall, 18 criteria were developed, clustered into four subcategories: I. Clinical criteria (characteristics of the target disease); II. Diagnostic criteria (requirements of the test); III. Therapeutic-interventional criteria (prerequisites of the intervention); IV. Program management criteria (requirements of the program). Subcategories I–III define selection criteria for target diseases, subcategory IV defines criteria for how to establish and manage the program.

**Conclusion:** This multi-dimensional framework serves as a well-balanced basis for developing thoroughly revised and internationally accepted consensus screening criteria.

## Introduction

Newborn screening (NBS) is among the most effective measures of secondary prevention and has helped to shift the traditional paradigm of medicine from symptom-driven to pre-symptomatic diagnosis in phenotypically healthy individuals.^1^ In parallel, genomic sequencing has been established as a routine healthcare component for individuals with symptoms of unknown etiology to guide treatment decisions and counselling of families.^2^

NBS is continuously expanded, with genomic sequencing expected to pave the next major expansion of this program.^3^ As information about all genetic susceptibilities is included in genomic datasets, a genomic NBS (gNBS) would allow for the detection of hundreds of target diseases currently undetectable with standard NBS techniques.^4,5^ To provide information only about target diseases, a virtual panel would have to be carefully set. Such a panel would allow rapidly including additional diseases and gene variants without extensive methodological effort, but would at the same time increase the risk of causing harm if diseases and gene variants were not carefully selected.

Decisions about which target diseases to include in NBS have long been guided by the Wilson and Jungner criteria.^6^ However, these criteria are insufficient to regulate a modern NBS program: they use imprecise terminology, lack measurability, a pediatric focus, and guidance on program management, leave a margin for interpretation, and are considered incomplete for the genomic approach.^7^ Adapted screening criteria have been proposed by various groups with only limited impact on (inter)national NBS regulations.^8–12^ Accordingly, the selection of target diseases for current NBS programs and gNBS pilot studies varies substantially.^13–15^ For example, among seven gNBS pilot studies with panels including 237 to 889 target diseases, only 53 target diseases overlapped, primarily representing inherited metabolic diseases already covered by current NBS programs.^14^

Thus, there is an urgent need to develop revised screening criteria before genomic sequencing can be effectively introduced into NBS programs. To achieve this goal, this study aims at developing a multidimensional approach through interdisciplinary discourse, integration of medical, ethical, legal, and social implications (ELSI), input from stakeholders as patient representatives, and consideration of program-management aspects.^1,7,16^

## Materials and Methods

To cover and integrate a broad range of relevant expertise for gNBS, an interdisciplinary expert panel was assembled, comprising experts from pediatric and adolescent medicine, human genetics, medical psychology, medical ethics, legal studies, and patient representatives, in form of the project group “NEW_LIVES: Genomic NEWborn Screening Programs – Legal Implications, Value, Ethics and Society”.^17^

First, each subdiscipline conducted and published extensive literature reviews.^7,14,18^ Additionally, ethical and legal implications were evaluated, and societal perspectives captured in three empirical studies: (1) A focus group study with adult patient representatives, parents of minors, and physicians from human genetics as well as pediatric and adolescent medicine,^19^ (2) a representative population-wide survey on attitudes toward gNBS, and (3) an online survey with parents exploring their views on gNBS. Second, based on these multidisciplinary results, the expert panel decided on a structure for potential screening criteria, consisting of four subcategories: (I) clinical, (II) diagnostic, (III) therapeutic, and (IV) program management criteria.^7^

A process similar to the Nominal Group Technique^20–22^ was used to reach consensus, gathering iterative feedback from all expert panel members and integrating it into the decision-making process (Figure 1). This process involved:

1. Initial suggestions for medical and ELSI considerations to be reflected in the screening criteria for gNBS informed by the results of the literature review,^7^ ethical and legal analyses, the empirical studies,^19^ and definitions of the ClinGen Gene Curation Working Group (ClinGen).^23^
2. Establishing a preliminary list of screening criteria for gNBS, which was continuously revised during intensive group discussions and revisions. This was based on structured feedback, also from international experts on gNBS during an Advisory Board meeting (January 2024, online) and an international conference on the topic (Heidelberg, Germany, March 2024).
3. Finalizing the wording of the screening criteria, and
4. Final approval by all group members (August 5, 2024).

**Figure 1.**
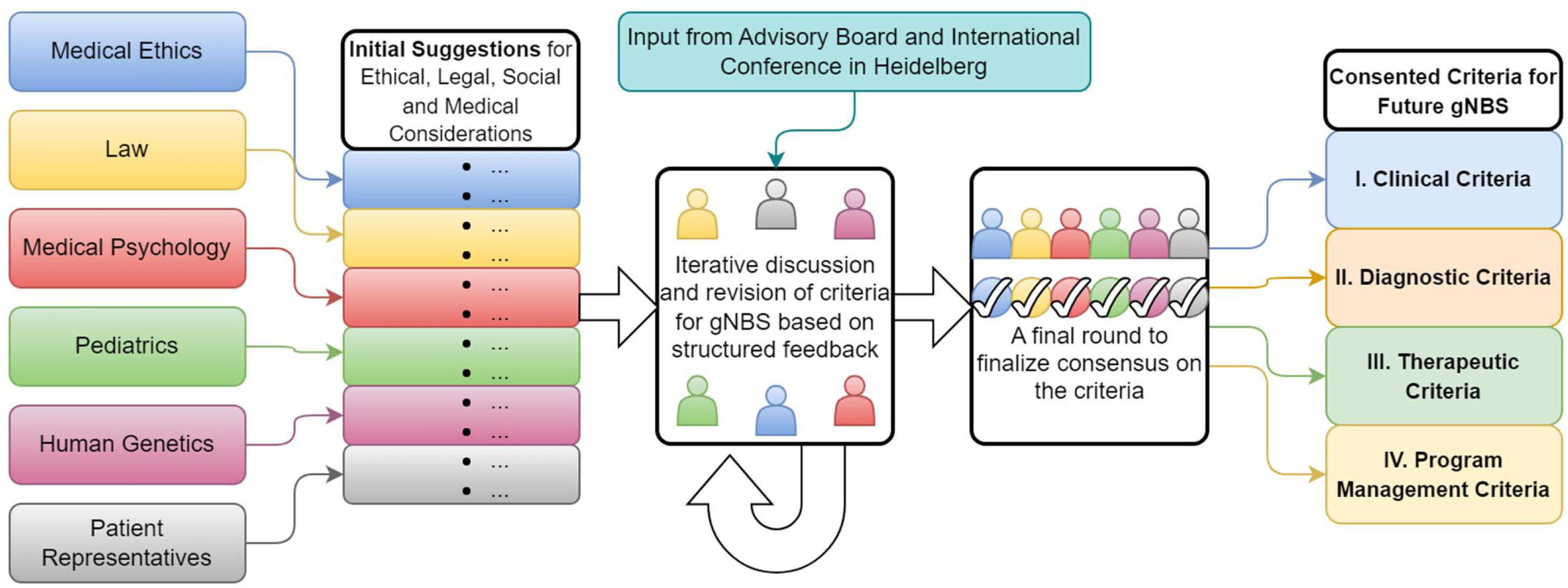
Working process for developing screening criteria for a future genomic newborn screening program. After initial suggestions regarding ethical, legal, social and medical considerations for potential screening criteria, a process similar to the Nominal Group Technique was applied to reach consensus decision, gathering iterative feedback from all experts, including international input from the advisory board and an international conference. The consented screening criteria are assigned to four subcategories: Clinical, Diagnostic, Therapeutic and Program Management Criteria. gNBS: genomic newborn screening. Figure was created with draw.io (https://drawio-app.com/, accessed on February 03, 2025).

## Results

The expert panel developed a multi-dimensional framework for a gNBS program consisting of 18 screening criteria (Table 1) that are assigned to two overarching categories (A and B) and four subcategories (I–IV):

A. Criteria that enable transparent disease selection:

I. 4 Clinical criteria,
II. 4 Diagnostic criteria, and
III. 3 Therapeutic-interventional criteria
B. Criteria to establish, manage, and further develop the gNBS program:

IV. 7 Program management criteria.

**Table 1.**
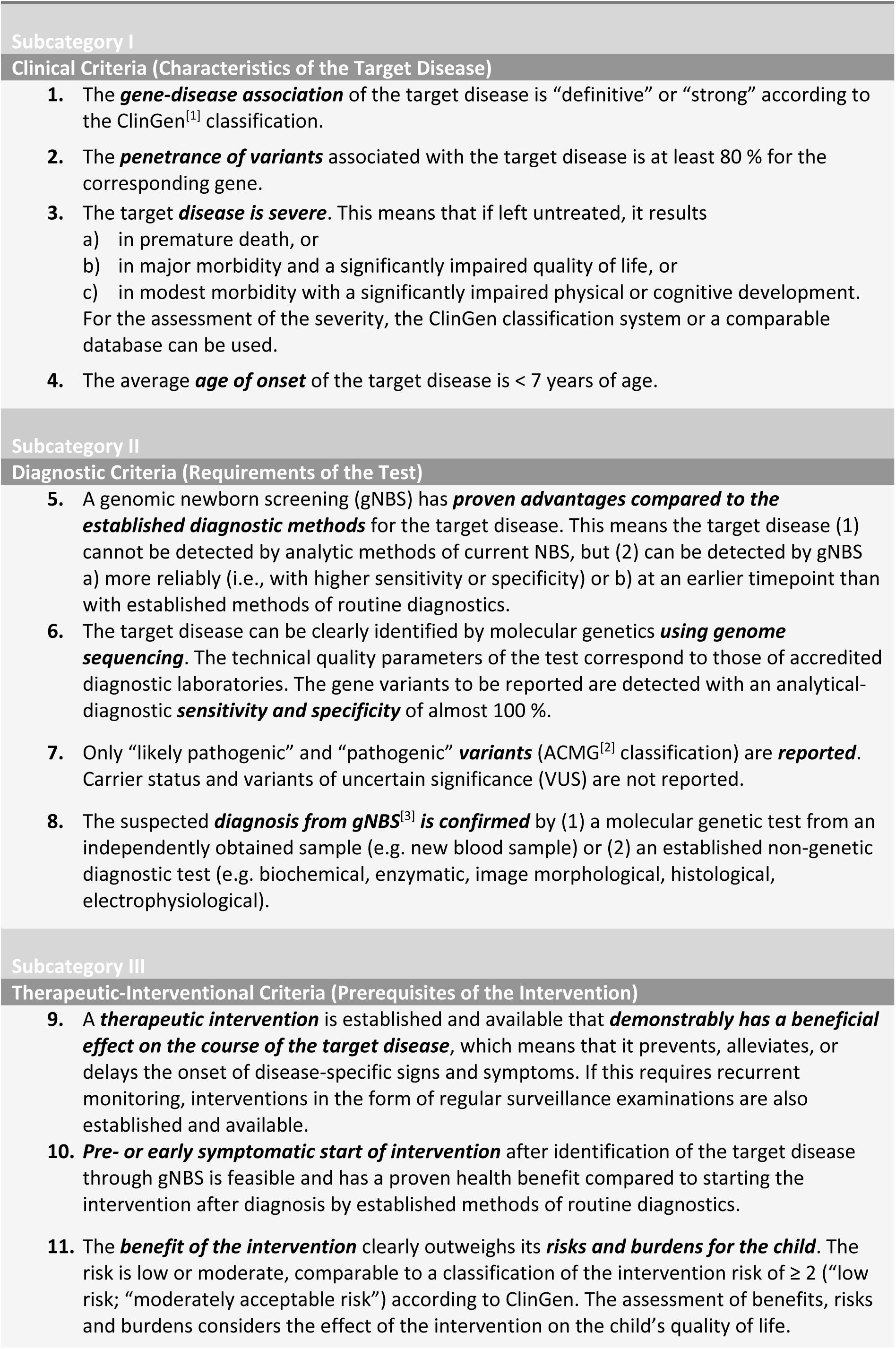
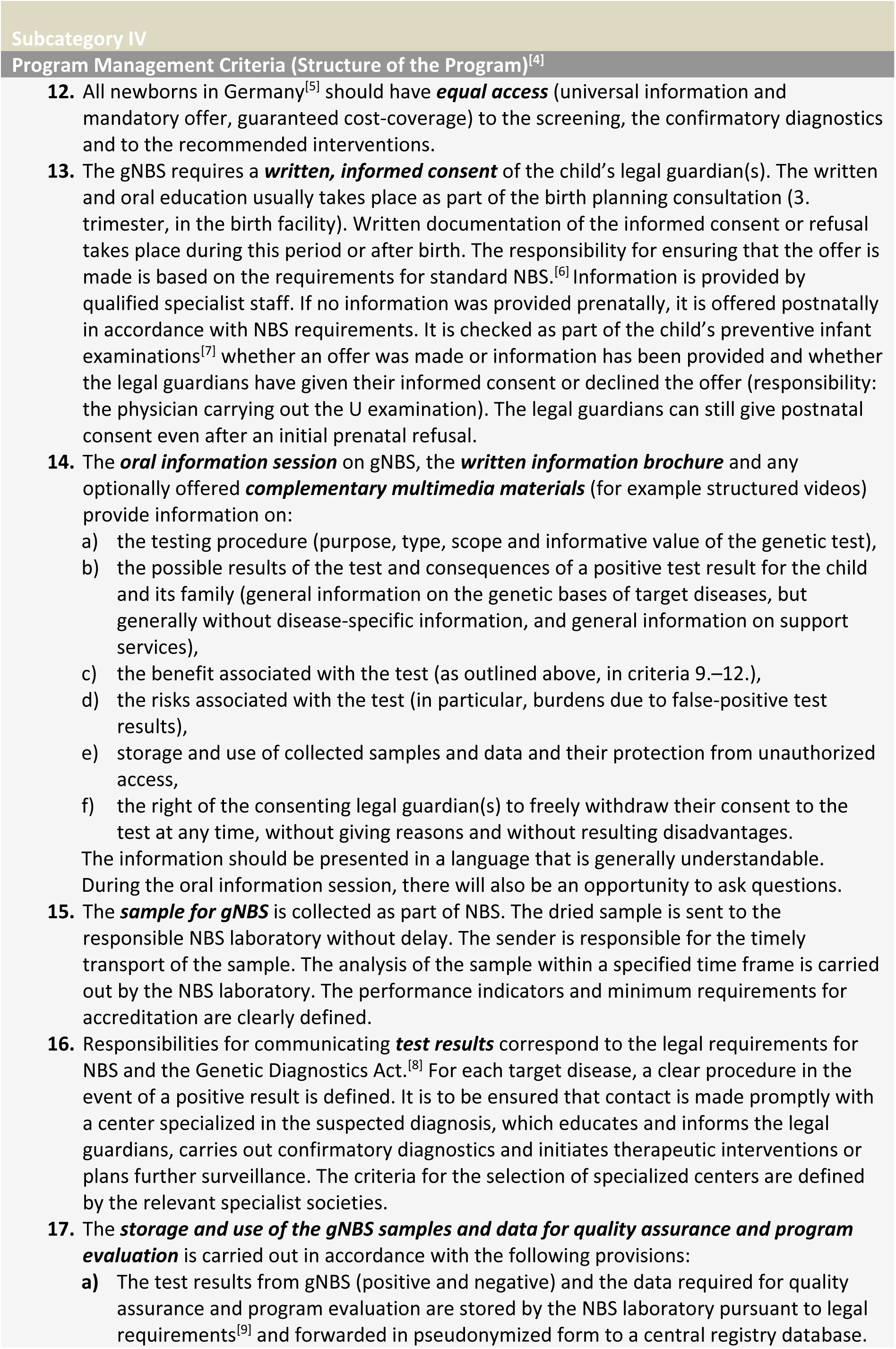
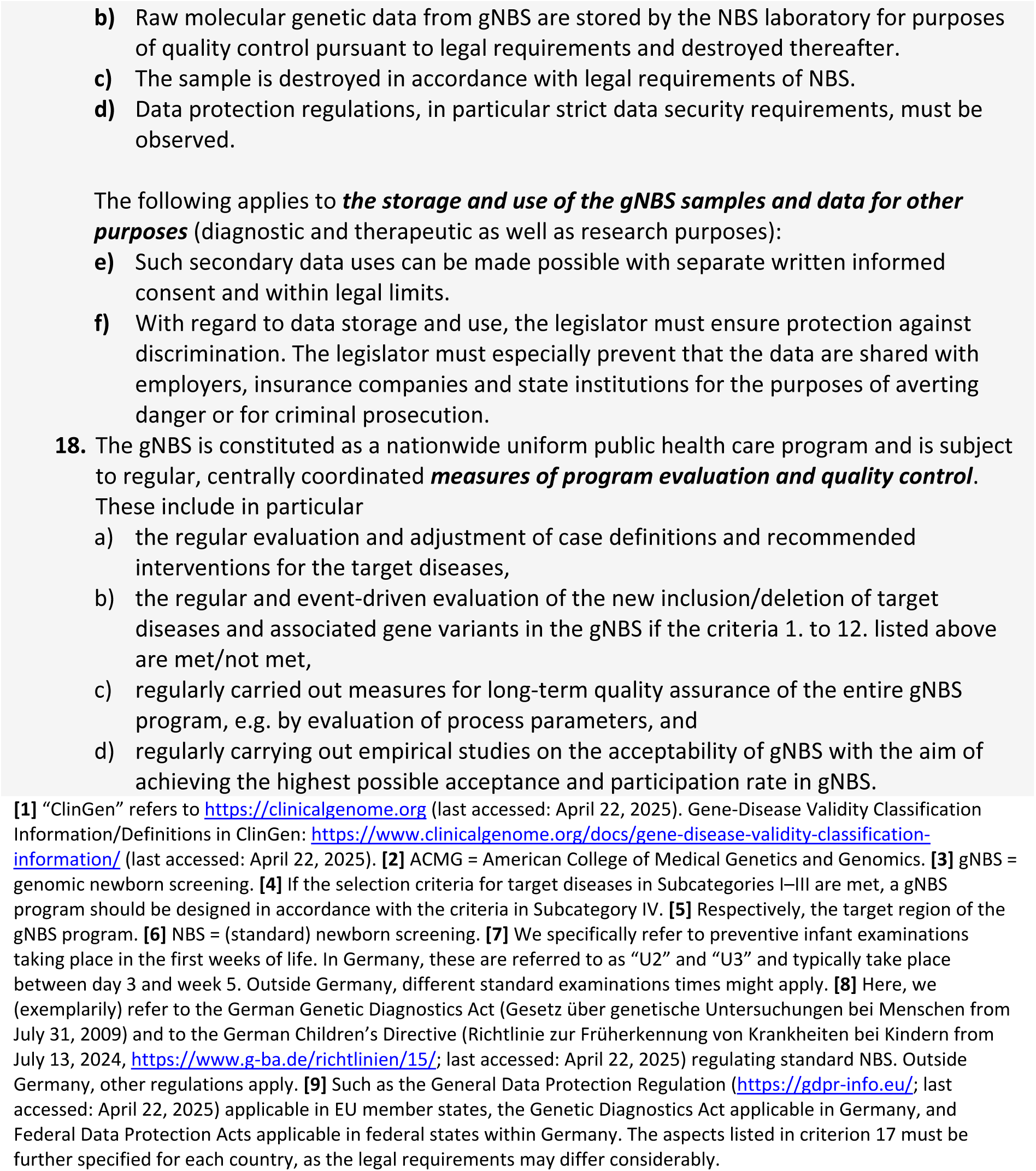
Criteria for a genomic newborn screening program (by the project group NEW_LIVES)

If all screening criteria in subcategories I–III are met, a target disease will qualify for inclusion in gNBS, while any (g)NBS program should meet all 7 management criteria of subcategory IV.

### I. Clinical criteria (Characteristics of the target disease)

1) The ***gene-disease association*** of the target disease is “definitive” or “strong” according to the ClinGen classification.

Evidence for the causal relationship between a gene and a disease should have been repeatedly demonstrated in both research and clinical diagnostics (“definitive”) or reported independently in ≥ 2 studies (“strong”). The expert panel recommends the use of a Gene-Disease Validity Classification Information, such as curated by ClinGen.^24^ For gene-disease associations not covered by ClinGen, the same standards as in ClinGen should apply.

2) The ***penetrance of variants*** associated with the target disease is at least 80 % for the corresponding gene.

Penetrance is the probability of the associated disease occurring in a person with a genetic variant.^25^ Penetrance is incompletely understood for many variants and target diseases.^26^ If a variant causes a genetic disease with multiple symptoms, the penetrance refers to the clinically relevant main symptom. Only variants or target diseases with high penetrance based on moderate to substantial evidence according to ClinGen^27^ should be considered. For target diseases not covered by ClinGen, the same standards as in ClinGen should apply.

3) The ***target disease is severe***. This means that if left untreated, it results

a) in premature death, or
b) in major morbidity and a significantly impaired quality of life, or
c) in modest morbidity with a significantly impaired physical or cognitive development. For the assessment of the severity, the ClinGen classification system or a comparable database can be used.

The expert panel has decided not to exclusively refer to the ClinGen classification as it leaves considerable room for interpretation. Diseases with “modest morbidity”, such as diseases leading to moderate intellectual disability, may not generally meet the definition of a severe disease. However, if associated with significant impairment of physical or cognitive development, they are considered severe (criterion 3 [#3]). In contrast, if modest or minimal morbidity without significant developmental impairment is commonly expected, #3 is not met and thus would not justify the inclusion in gNBS.

4) The average ***age of onset*** of the target disease is < 7 years of age.

To avoid waiting periods until disease onset and as applicable for most target diseases of current NBS programs, only diseases with an average age of onset before school age should be included. As genetic diseases usually have a wide range of age of onset, the average should always be considered, which is justified by a trade-off between the benefit for children with earlier onset and avoidance of “patients in waiting”.^28^ This is particularly important as any inclusion of a disease into NBS will lead to expansion of the known clinical phenotype, especially towards attenuated phenotypes, which are likely to shift the age of onset to a later timepoint.^1^

### II. Diagnostic criteria (Requirements of the test)

5) A gNBS has ***proven advantages compared to the established diagnostic methods*** for the target disease. This means the target disease (1) cannot be detected by analytic methods of current NBS, but (2) can be detected by gNBS a) more reliably (i.e., with higher sensitivity or specificity) or b) at an earlier timepoint than with established methods of routine diagnostics.

A gNBS should have diagnostic advantages compared to both NBS (#5 (1)) and symptom-based routine diagnostic methods (#5 (2)). Consequently, gNBS is not intended to replace current NBS but to be added for diseases that cannot be identified by current screening methods. For these diseases, gNBS must have the above-mentioned (#5 (2)) advantages for detecting the target disease compared to routine diagnostics in a symptomatic child.

6) The target disease can be clearly identified by molecular genetics ***using genome sequencing***. The technical quality parameters of the test correspond to those of accredited diagnostic laboratories. The gene variants to be reported are detected with an analytical-diagnostic ***sensitivity and specificity*** of almost 100 %.

The quality parameters of the test must meet the highest diagnostic standards and (likely) pathogenic variants in a given sample must be detected. However, incomplete clinical sensitivity, caused by a high proportion of variants of uncertain significance (VUS), some of which may later be classified as pathogenic, should not be considered a general exclusion criterion for a target disease: if not all affected individuals receive a genetic diagnosis, those with a (likely) pathogenic variant in this gene would still benefit from reporting the findings. Genome sequencing can be used to identify single nucleotide variants, small rearrangements, and copy number variations, while e.g. methylation defects cannot be identified with the genome-sequencing methods commonly used today. This must be considered when selecting target diseases and adapted to new technologies.

7) Only “likely pathogenic” and “pathogenic” ***variants*** (ACMG classification) are ***reported***. Carrier status and variants of uncertain significance (VUS) are not reported.

The American College of Medical Genetics and Genomics (ACMG) criteria for interpretation of sequence variants recommend five categories for classifying the probability that the variant is disease causing: pathogenic, likely pathogenic, uncertain significance (VUS), likely benign, and benign.^29^ Information on symptoms and family history is required for classification, which is not available in the screening context. Therefore, and to achieve the highest possible specificity, only variants of the first two categories should be reported. Autosomal recessive (AR) heterozygous variants not relevant for the health of the newborn should not be reported: in AR inheritance, an individual is heterozygous if only one of the two gene copies carries a (likely) pathogenic variant, whereas the full clinical picture would only occur if both gene copies were altered. In X-linked inheritance, hemizygous males (karyotype 46, XY) are affected by the disease, whereas heterozygous females (46, XX) are usually less severely affected or unaffected; exceptions exist. Therefore, (likely) pathogenic variants in target diseases with X-linked inheritance are usually only reported in males and not in heterozygous females.

8) The suspected ***diagnosis from gNBS is confirmed*** by (1) a molecular genetic test from an independently obtained sample (e.g. new blood sample) or (2) an established non-genetic diagnostic test (e.g. biochemical, enzymatic, image morphological, histological, electrophysiological).

Screening tests differ from diagnostic tests as they identify individuals at risk for the identified condition preferably during the preclinical stage of the disease. Thus, positive screening tests always require a careful and reliable confirmatory strategy, particularly, as in rare diseases false positive results are relatively frequent. For current NBS, biochemical and targeted genetic tests are used for confirmation.^30^ Target diseases included in gNBS programs equally require a predefined pathway of suitable confirmatory tests, such as non-genetic functional biomarkers. They can be based on a secondary genetic analysis and an alternative molecular genetic test (e.g. Sanger sequencing) for the suspected disease in a new sample if non-genetic tests are not established. A careful evaluation of the phenotype and family history should be used to complete the confirmatory pathway. If the confirmatory diagnostics are based on a second genetic test, the biallelic phasing of the variants should be confirmed in AR inheritance. If there are medically equivalent ways of confirmatory diagnostics, the one with the least discomfort for the child should be chosen.

### III. Therapeutic-interventional criteria (Prerequisites of the intervention)

9) A ***therapeutic intervention*** is established and available that ***demonstrably has a beneficial effect on the course of the target disease***, which means that it prevents, alleviates, or delays the onset of disease-specific signs and symptoms. If this requires recurrent monitoring, interventions in the form of regular surveillance examinations are also established and available.

Other possible benefits, such as non-medical actionability, secondary benefits to parents, siblings, or society, family planning implications, avoidance of a “diagnostic odyssey”, or just the wish to know, are not considered sufficient to justify the inclusion of a target disease in a population-wide NBS or gNBS.

An ongoing clinical trial would not suffice to consider an intervention as “established”.

“Available” means that all screened individuals have equal access to treatment and to specialized teams offering monitoring and treatment. If gNBS also included diseases that did not require therapeutic intervention from the time of diagnosis but recurrent surveillance, e.g. *RB1*-related retinoblastoma, serial examinations necessary for regular surveillance as well as therapeutic interventions in the event of disease development would have to be established and made available as defined above.

10) ***Pre- or early symptomatic start of intervention*** after identification of the target disease through gNBS is feasible and has a proven health benefit compared to starting the intervention after diagnosis by established methods of routine diagnostics.

If pre- or early symptomatic initiation of therapy did not provide a health benefit, post-symptomatic targeted routine diagnostics would be sufficient and the candidate disease would not qualify for inclusion as target disease in gNBS. Since genome sequencing is frequently performed at the appearance of first symptoms and even ultra-rapid genome sequencing is increasingly available for critically ill children, the sole prevention of a “diagnostic odyssey” does not justify the inclusion into gNBS.^31^

11) The ***benefit of the intervention*** clearly outweighs its ***risks and burdens for the child***. The risk is low or moderate, comparable to a classification of the intervention risk of ≥ 2 (“low risk”; “moderately acceptable risk”) according to ClinGen. The assessment of benefits, risks and burdens considers the effects of the intervention on the child’s quality of life.

The known risk and burden of therapeutic interventions or surveillance examinations should be outweighed by their potential benefit. Since the assessment of risks and benefits includes normative components, patient representatives should be involved in the process alongside medical and medical-ethics experts. Priority should always be given to the quality of life of the affected child because the burden and risk of an intervention as classified in ClinGen^32^ may not (always) correspond to the perception of those affected.^33^

### IV. Program management criteria (structure of the program)

Any NBS, including a gNBS, should be regarded as a public health measure. To offer guidance for the process of establishing, evaluating, and maintaining a (g)NBS program, seven criteria for program management have been defined (##12–18). For detailed criteria, see Table 1, key considerations are summarized below.

***Equal access* (#12)**, which is an important ethical requirement of any public health program, comprises the guarantee to cover costs of screening, confirmatory diagnostics, and consecutive interventions and care. The current NBS programs generally provide equal access to all newborns in the target region and thus enable a very high participation rate, e.g. about 99.9 % in Germany.^34^ In contrast, cost coverage for recommended therapies is still incomplete, e.g. special diets are not regularly paid for by health insurances.^35^ Nevertheless, achieving the greatest possible equality, including full-cost coverage of recommended follow-up care, should be the aim of any NBS program.

While equality of access (#12) implies that every newborn’s guardians are offered gNBS, ***participation needs to remain voluntary* (##13–14)**. Although written informed consent (IC) is not uniformly required across all NBS programs worldwide,^36^ it will be mandatory for gNBS as for any genetic diagnostic. Since gNBS is a complex topic, there is a balance to be achieved between information overload and insufficient information. Hence, the expert panel specifies essential content in #14a–f that should be addressed. Withdrawal from gNBS is possible at any time (#14f) with the exception that a positive gNBS result, once obtained, will be communicated in the interest of the child in any case.

To make gNBS as minimally invasive for the child as possible, the ***same dried blood spot sample*** should be used for NBS and gNBS, utilizing the established system of filter paper cards. Sample collection, transport, and analysis **(#15)**, confirmation and communication of test results, and the initiation of recommended interventions **(#16)** should follow clearly defined guidelines including exact procedures for each target disease. It is essential for a gNBS program that precise information about the procedure following a positive result, including specific contact persons, is provided to families directly after a positive screening report.

The use of data and samples from gNBS requires ***strict adherence to data protection laws***, including national and international regulations **(#17)**. Samples and data from gNBS are of great value for program evaluation and quality assurance (#17a–d), but also for adjustment and improvement of the gNBS program. In addition, they might contribute to research on genetic diseases potentially resulting in new or improved treatment, and at the individual level. Since genetic data is considered particularly sensitive, it requires special protection. Therefore, the expert panel recommends that such secondary use of data should only be made possible either with separate written IC or where legally permitted (#17e), in both cases with additional safeguards (#17f), in compliance with applicable law and regularly updated to reflect changing legal requirements.

***A gNBS should be organized as an integrated and learning public health program*** with central coordination, data-driven evaluation of quality, safety, and acceptability, and re-evaluation of target diseases, including case definitions and recommended interventions **(#18)**.

## Discussion

The proposed screening criteria aim to establish a transparent multi-dimensional framework for the selection of target diseases (Table 1, I–III) and the management of gNBS programs (Table 1, IV). They should guide the future development of NBS programs in general but have a specific focus on gNBS as the expected next major NBS expansion.

### Towards harmonization of selection criteria for target diseases

Currently, the selection of target diseases for international NBS programs and gNBS pilot studies varies considerably.^13–15^ Several factors may contribute, including country-specific priorities, different medical backgrounds of pilot project investigators, and differently weighted screening criteria. For instance, NC NEXUS focused on actionability,^5^ while BabySeq prioritizes diagnostic certainty with high penetrance and high gene-disease validity.^4^ The overarching question is how screening criteria should be designed to lead to sufficient agreement and international harmonization. This is particularly important as, without consensus, there is a high risk of increasing confusion and variability in target disease-lists. At the same time, target lists need objective criteria to stay flexible amid future innovation and gene-disease connections, not yet identified. A model for unifying different screening panels might be the Recommended Uniform Screening Panel in the United States.^37^ Our proposed screening criteria offer a potential first step toward a promising approach: to initially apply conservative and stringent screening criteria for defining a core gNBS panel, ensuring consistency and clarity in its development.

### A balanced approach to selecting screening criteria

The proposed screening criteria are designed conservatively to avoid uncertainty and to improve acceptability by the following three measures:

1. Each of the criteria must be fulfilled, and all criteria are treated equally. While alternative approaches like a prioritization with major and minor criteria, or a scoring system would be conceivable,^5,38^ the weighing would not be evidence-based and target diseases could enter the list with questionable benefit – which is why we recommend to apply all of the 18 criteria.
2. The screening criteria focus on the benefit and best interests of the newborn. Potential benefits to parents or siblings alone are insufficient for the inclusion as a target disease.
3. The criteria themselves are designed to be conservative and stringent. For example, penetrance must be above 80 %, the disease must occur on average (and not at the earliest) before the age of 7 years, and not only a therapeutic benefit, but also the benefit of a pre-symptomatic start of intervention, must be proven.

There are (a) medical, (b) ethical, (c) legal, and (d) societal and psychological reasons supporting the conservative approach.

a. NBS is a predictive test for individuals who are considered healthy and have a cumulatively low risk of being affected by an NBS target disease. There are no symptoms to guide the interpretation of test results. For parents, the primary expectation of NBS is not the diagnosis of a condition, but rather the confirmation of their child’s healthiness. Therefore, it is crucial that conditions are detected with the highest possible certainty, that a reported condition will manifest with high likelihood, and that an effective intervention can be provided. Unclear findings, including conditions with low penetrance or late-onset conditions may cause significant anxiety. Uncertainty may lead to overdiagnosis and overtreatment as well as rejection of the program.
b. The proposed conservative approach for the framework for gNBS described here is supported by the four core principles of biomedical ethics: nonmaleficence, beneficence, justice, and respect for autonomy.^39^ Stringent requirements for, e.g., gene-disease association (#1), penetrance (#2), variant pathogenicity (#7), and confirmation of suspected diagnoses (#8), serve the purpose of preventing harm for newborns and their parents by reducing the number of “patients in waiting”^28^ and “parents in waiting”. The primary benefit of gNBS to prevent severe genetic diseases can only be guaranteed – and guaranteed justly – if stringent requirements for sensitivity and specificity of the proposed screening methods apply (#6), and if therapeutic interventions and any recommended additional surveillance interventions are available (#9) as well as equally accessible (#12). The stringent requirements for the IC process (##13–14, 17) are ethically justified by the principle of respect for autonomy.
c. The conservative approach can also be supported by legal arguments pertaining to the German context, e.g. the specifications for the content of the IC session as described in #14 a–f largely reflect the requirements as specified in § 9 para 2 no 1–6 in the German Genetic Diagnostics Act (GenDG). Similarly, the requirement in #9 that a therapeutic intervention must be established and available for the target condition is consistent with the provisions of § 16 para 1 GenDG. Furthermore, the requirement in #11 that the benefits of the intervention must clearly outweigh the risks and burdens for the child ensures compliance with the fundamental principle of the best interests of the child.
d. The proposed stringent requirements, e.g. for high penetrance (# 2), data protection (#17), IC (##13–14), and focus on severe diseases (#3), are intended to promote broad societal acceptance of gNBS (see also #18d). Such acceptance is considered crucial for the sustainable implementation and broad participation in the program. Previous empirical studies identified concerns regarding data protection and privacy,^19,40,41^ as well as the uncertainty surrounding results that lack clear clinical implications.^19,41–43^ The potential for emotional distress and psychological burden arising from the ambiguity of findings in population genetic screening has also been emphasized,^44^ supporting a more cautious and restrictive approach also from a psychological perspective. While several studies have shown that parents express interest in a broad spectrum of information—even in the absence of actionable treatments—there is nevertheless a stronger consensus in favor of disclosing findings related to conditions with specific therapeutic options.^45–47^ Finally, the BabySeq project revealed greater support for the existing NBS than for gNBS, suggesting a general inclination toward more stringent inclusion criteria.^45^

### Proposed approach – Possible limitations and unresolved concerns

The proposed screening criteria focus on severe, early onset and treatable target diseases to reduce uncertainty and maximize the benefit for newborns. In contrast to this, parents and clinicians sometimes highlight genomic data’s personal utility for families even in the absence of medical utility for the child.^15,48^ In this context, certainty of prediction seems to be assigned higher priority than treatability,^49^ and benefits may be expected for “family planning and testing, the intrinsic value of information, and the ability to prepare for the future.”^48^ For these reasons, some gNBS pilot projects offer a complementary opt-in for extended categories of results.^4,15^ This would offer parents more autonomy in deciding on the return of results than suggested here. There are, however, also indications that parents have exaggerated expectations of gNBS and overestimate its potential benefits.^48^ This should be considered when developing selection options and IC materials, and a strict analogy between gNBS pilot projects and a population-wide gNBS program, as discussed in this study, cannot be drawn.

Another possible disadvantage of a conservative approach may be that genetic information is not reported if screening criteria are not all met but might still have a proven medical benefit. For example, not reporting VUS is an internationally recognized approach for gNBS;^4^ however, this reduces sensitivity (for the sake of specificity and positive prediction), as VUS could otherwise be re-evaluated later or in a family context.

Precise knowledge of the genetic variants in the screened population is an important condition for the success of a gNBS. A particular challenge here is achieving equity for minorities, for whom only limited population-specific genetic data might be available for genomic analysis. As with any screening, a negative gNBS result does not exclude a hereditary disease for methodological and scientific reasons such as incomplete knowledge about elements of gene regulation. Therefore, gNBS cannot replace symptom-related genomic diagnostics in the clinical context. If symptoms occur that are thought to have a genetic cause, a clinical genetic evaluation must still be carried out.

Due to the complexity and limitations, the development of IC materials, possibly supported by chatbots and videos, should be given high importance in concrete program development. The associated costs for the development of such materials, as well as for the education and training of health professionals ensuring a high-quality IC process must be included in any future health economic assessment of gNBS.

### Program management, centralized data collection, and learning system

For the continuous optimization of future gNBS programs, a gNBS should ideally be designed as an integrated public health and learning healthcare program with centralized structures for data collection and the establishment of a registry. Regular analysis of key performance indicators is essential for data-driven evaluation and continuous optimization of the program, ideally supported by explainable AI-based algorithms. This is particularly important because changes in the knowledge about natural history (e.g. phenotype diversity) or the establishment of new therapeutic interventions may change the achievement of screening criteria for a target disease and, therefore, the disease may be added to or removed from gNBS. In fact, a negative evaluation should enable the removal of target diseases from the gNBS program through a previously defined transparent mechanism. In addition to the core panel, pilot studies may be conducted to assess technical feasibility and health benefits. Together with registry-based evaluations, results of pilot studies may lead to future re-assessment of individual criteria.

## Conclusion

The multi-dimensional framework presented here has been systematically developed by an expert panel representing medicine, ELSI and patient representatives. It includes 11 selection criteria for target diseases and 7 program management criteria for future gNBS programs. By applying a deliberately stringent and child-centered set of criteria, the framework aims to achieve high acceptance and to facilitate international harmonization of screening criteria. Additionally, it could serve as a well-balanced basis for the harmonization of a core-panel of target diseases for gNBS.

## Abbreviations

ACMG: American College of Medical Genetics and Genomics
AR: Autosomal Recessive
ClinGen: ClinGen Gene Curation Working Group
ELSI: Ethical, Legal, and Social Implications
GenDG: German Genetic Diagnostics Act (Gendiagnostikgesetz)
gNBS: Genomic Newborn Screening
IC: Informed Consent
NBS: Newborn Screening
VUS: Variant of Uncertain Significance

## Data availability

not applicable.

## Acknowledgements

We thank all colleagues of the NEW_LIVES Advisory Board and all contributors at the NEW_LIVES conference in Heidelberg for their intensive discussion and valuable contributions. Additionally, we thank our colleagues from the Institute for Medical and Data Ethics, previously Section Translational Medical Ethics (AG Winkler), the Institute for Human Genetics, the Section for Pediatric Neurology and Metabolic Medicine and the Newborn Screening Laboratory at Heidelberg University and Medical Faculty of Heidelberg University Hospital, as well as attendees of meetings and conferences where these criteria were presented, for their valuable feedback.

## Author Contributions (CRediT)

Conceptualization: E.S.-B., N.D., K.A., S.S., B.D., R.M.-T., C.P.S., S.K., E.W.; Funding acquisition: B.D., R.M.-T., C.P.S., S.K., E.W.; Investigation: E.S.-B., N.D, K.A., U.M., H.S., E.S.D., J.M., H.B., C.J.M., L.N., T.H., H.H., S.S., B.D., R.M.-T., C.P.S., S.K., E.W.; Project administration: K.A., L.N., S.S., B.D., R.M.-T., C.P.S., S.K., E.W.; Resources: B.D., R.M.-T., C.P.S., S.K., E.W.; Supervision: B.D., R.M.-T., C.P.S., S.K., E.W.; Visualization: E.S.-B., N.D, K.A., L.N., C.P.S., S.K., E.W.; Writing-original draft: E.S.-B., N.D, K.A.; Writing-review & editing: U.M., H.S., E.S.D., J.M., H.B., C.J.M., L.N., T.H., H.H., S.S., B.D., R.M.-T., C.P.S., S.K., E.W.

## Ethics Declaration

For this article no studies with human or animal subjects were performed. All procedures followed were in accordance with the Declaration of Helsinki.

